# Comparison of High-Resolution Computed Tomography Patterns in Adult with Cystic Fibrosis and Non-Cystic Fibrosis Bronchiectasis in a South Asian Country Bangladesh: A Retrospective Cross-sectional Study

**DOI:** 10.64898/2026.03.09.26347994

**Authors:** Shuvo Majumder, Pujaneeta Biswas, Rajashish Chakrabortty, Shamim Ahmed, Mohammed Atiqur Rahman

**Affiliations:** Department of Respiratory Medicine, Bangladesh Medical University, Dhaka, Bangladesh; Department of Laboratory Medicine, Bangladesh Medical University, Dhaka, Bangladesh

**Keywords:** Chest HRCT, bronchiectasis, CF, non-CF

## Abstract

**Background:** Bronchiectasis in adults often goes undiagnosed following the routine assessment. Cystic Fibrosis (CF) is usually diagnosed during childhood, but some cases are identified in adulthood when disease is mild. High-resolution computed tomography (HRCT) of chest may offer structural information that can indicate CF as an underlying etiology.

**Objective:** To compare the HRCT features of adult patients with CF and non-CF bronchiectasis and to determine the radiologic features that may be suggestive of CF.

**Methods:** This retrospective, analytical, cross-sectional study was carried out in Bangladesh Medical University after IRB clearance. Total 130 adults (12 with CF and 118 with non-CF bronchiectasis) of both sexes, whose bronchiectasis was confirmed by chest HRCT were included. Imaging findings were assessed based on Reid morphological classification, anatomical distribution and extent of spread within the lungs, and their association was tested using chi-square test with statistical significance of p<0.05.

**Results:** Cystic bronchiectasis was more common in CF than non-CF patients (83.3% vs 29.7; p<0.001). Mixed central-peripheral extension had been found a considerable associated with CF (66.7% vs. 42.4; p=0.034). There was no statistically significant difference in right lung lobar distribution (p=0.540) but combined upper and lower lobe involvement on the left side was more common in CF patients (54.5%) than non-CF patients (21.3) (p=0.054).

**Conclusion:** Adult CF had unique chest HRCT imaging characteristics when compared to non-CF bronchiectasis, especially cystic morphology and mixed extension. Identification of such features could help physician in the early diagnosis and selection of treatment strategy.

## Introduction

Bronchiectasis is a chronic respiratory disease that is associated with permanent abnormal dilatation of the proximal sub-segmental bronchi and is typically manifested by coughing, daily sputum production, and frequent lower respiratory infection with varying levels of airflow obstruction^1^. Etiologies in adult populations are usually post-infectious damages, pulmonary tuberculosis, immunodeficiency, and hypersensitivity disorders; however, a certain percentage of patients do not have a well-defined cause following routine examination^2^. Bronchiectasis is a highly prevalent comorbid condition in Asia and the actual prevalence of bronchiectasis in Asia is unknown due to limited studies. According to the Indian bronchiectasis registry, idiopathic is the most prevalent cause of bronchiectasis, although post-TB bronchiectasis is more prevalent^3^. Bronchiectasis predisposes to atypical pulmonary infections and hemoptysis, caused by angiogenesis and hypertrophy of bronchial arteries induced by inflammation^4^.

Cystic fibrosis (CF) is a genetic disease that results because of the dysfunction of the CF transmembrane conductance regulator (CFTR) protein^5^. Advances in awareness and diagnostic capacity have shown that a few CF patients appear in adulthood with only respiratory defects, such as bronchiectasis^5^.

Bronchiectasis places a huge burden on patients regarding symptoms, lung function, exacerbations, and quality of life. Genetic counseling and early application of treatment can improve the quality of life and even lifespan of affected individuals by detecting CF at an early stage. HRCT may assist in detecting CF as it can identify the characteristic patterns early in the disease progression that can aid the physician to initiate the appropriate measures for managing the disease, since genetic testing is unavailable due to scarce resources to confirm the disease.

High-resolution computed tomography (HRCT) of chest is the most ideal imaging tool to assess structural airways^6^. Some patterns, especially the presence of advanced cystic changes and the upper lobe predominance, can indicate CF despite having low clinical suspicion^7^. Mosaic attenuation, secondary to air trapping, is the main CT finding during the early stages of cystic fibrosis (CF). Bilateral bronchiectasis, which is mostly in the upper lobes, is other typical CT features, while the lower lobes tend to be affected in non-CF conditions^8^. Apparently, early lung disease observed during CT scans is characterized by the air trapping and thickening of the airway walls, dilation of the airways is both signs of active disease progression, whereas development of permanent bronchiectasis is a terminal outcome caused by prior pathological destruction^9^. In adult patients with cystic fibrosis, radiographic changes in HRCT shows a faster progression than indices of spirometry. When CT scoring is corelated with lung function, peripheral bronchiectasis CT score decreases at a faster, more frequent rate (1.52% per year) than lung function measures in adults with cystic fibrosis. In addition to this, deterioration of certain HRCT features depends on the extent of compromise of underlying lung functions^10^. The latest developments have increased the number of clinical uses of CT in bronchiectasis, and it has been used more frequently into diagnostic and prognostic assessment guidelines to measure treatment response^11^. Recent clinical practice guidelines by the Cystic Fibrosis Foundation suggest that CT scans should be done every 2-3 years with lowest possible radiation dose^12^. A cohort study carried out in the Netherlands indicating that pulmonary functions testing and CT imaging measures various aspects of CF-related lung disease^13^. The purpose of the given study was to describe the features of the HRCT of chest in patients with bronchiectasis, as well as to compare the radiological patterns in between CF and non-CF etiology. This study also aimed at establishing some distinguishable imaging characteristics that can be used to differentiate CF-related bronchiectasis against other common etiologies, especially where post-tubercular bronchiectasis is high-burden regarding this context like Bangladesh.

## Materials and Methods

The present retrospective cross-sectional analytical study was conducted in the Department of Respiratory Medicine at Bangladesh Medical University (BMU), Dhaka, Bangladesh, based on the data collected on a thesis research project that was completed previously between January 2023 and December 2024. Adult patients (aged 18 years and above) of both sex presenting with known bronchiectasis effecting one or more lobes in the high-resolution computed tomography (HRCT) were included. Non-probability consecutive sampling technique was done and the sample size was calculated by a formula to compare two proportions that led to a sample size of 130 participants. Those with traction bronchiectasis due to interstitial lung disease, post-tuberculosis bronchiectasis, or those who could not or did not wish to give informed consent were excluded. The data was derived on the basis of the archived data of the original thesis database and analyzed secondary on 7^th^ of January in 2026. Diagnosis of cystic fibrosis (CF) was performed on the basis of sweat chloride test, and patients were grouped into CF and non-CF bronchiectasis. An experienced radiologist who did not have access to clinical data evaluated HRCT images and categorized bronchiectasis by Reid morphology (cylindrical, varicose, and cystic), and lobar distribution and extent of lung involvement (central, peripheral, or mixed). The analytical work was done in SPSS version 25.0 (IBM Corp., Armonk, NY, USA). The chi-square test was used to compare categorical variables and a p-value of less than 0.05 was taken as statistically significant. The study received ethical consent of the Institutional Review Board of BMU, and all information was anonymized; the authors did not obtain identifiable participant information before or after extraction and analysis of data.

## Results

A total of 130 known adult bronchiectasis (12 CF and 118 non -CF) patients aged 18 years or older were included in this study and HCRT of chest imaging was analyzed according to study objectives. The morphological classification (Reid Type) of bronchiectasis in Fig 1 indicated that cystic bronchiectasis was the most common type in CF patients (83.3%), then cylindrical (8.3%), and varicose (8.3%), whereas in non-CF patients, the most common type was cylindrical (44.1%), then cystic (29.7%) and then varicose (26.3%); (p-value = <0.001).

**Fig-1:**
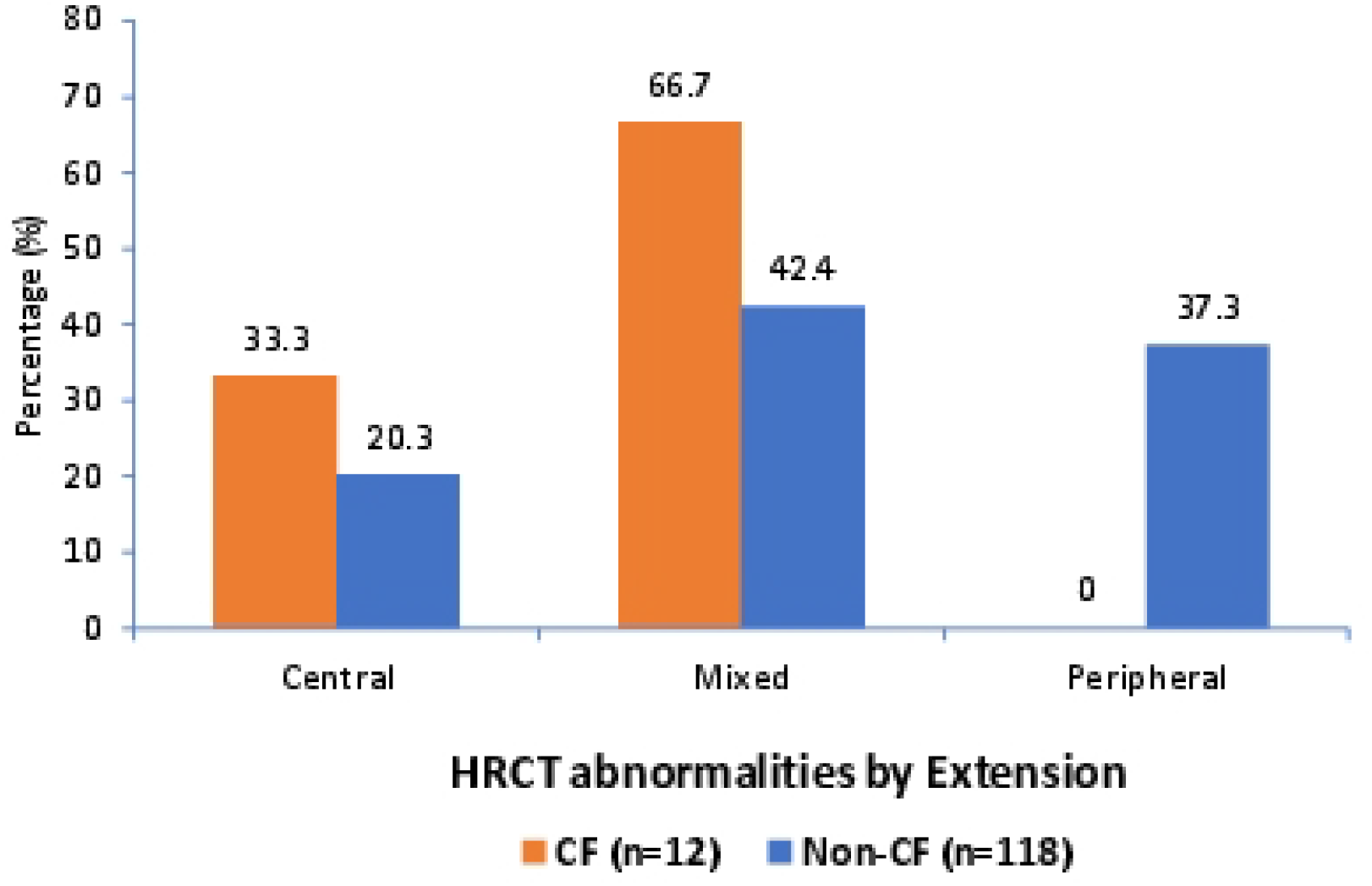
Bar diagram showing comparison of HRCT abnormality according to extension of bronchiectasis within the lung (N=130).

**Table 1.** Bar diagram showing the comparison of HRCT abnormality according to morphology of bronchiectasis (N=130). Table 1 Legend: Cystic bronchiectasis was much more common in CF patients (83.3) than in non-CF patients (29.7) (p = 0.001), but cylindrical bronchiectasis was more common in the non-CF group (44.1%).

| Reid morphology of bronchiectasis | CF<br>(n=12) ‡ |  | Non – CF<br>(n=118) ‡ |  | p-value* |
| --- | --- | --- | --- | --- | --- |
|  | N | % | n | % |  |
| Cylindrical | 1 | 8.3% | 52 | 44.1% | <0.001 |
| Varicose | 1 | 8.3% | 31 | 26.3% |  |
| Cystic | 10 | 83.3% | 35 | 29.7% |  |
| Total | 12 | 100.0% | 118 | 100.0% |  |

‡ n = number
\*p-value obtained by Chi-square test, $p < 0.05$ was considered as a level of significant

When extension patterns of bronchiectasis were assessed, it revealed that in CF patients, the distribution of bronchiectasis was mostly mixed (66.7%) and isolated peripheral involvement was not identified. On the contrary, the non-CF group showed more prevalence of peripheral bronchiectasis (37.3%) apart from mixed (42.4%) patterns; (p-value = 0.034), displayed in Fig-1.

CF patients had more mixed and central bronchiectasis, whereas peripheral bronchiectasis was found only in the non-CF patients.

Lobar distribution of bronchiectasis in Table 2 showed that, on the right lung, there was no significant difference in the distribution of HRCT abnormalities between CF and non-CF patients (p-value = 0.540). Combination of middle and lower lobe involvement was the most frequent in the CF group (n = 12, 33.3%), and then, there was isolated middle lobe involvement (25.0%). In the non-CF group (n = 106), combined upper and middle lobe involvement was common (20.8%), followed by the isolated middle (19.8%) and lower lobe involvement (17.0%). A borderline difference was observed on the left lung (p-value = 0.054). In CF patients, combined upper and lower lobe involvement was most common (54.5%), and isolated upper lobe involvement was most common in non-CF patients (52.8%).

**Table 2:** Comparison of HRCT abnormality according to lobar distribution within the lungs (N=130) Table 2 legend: There was no statistically significant difference in right lung involvement (p = 0.540). But the combined upper and lower lobe disease was more common in CF patients (54.5) compared to the non-CF patients (21.3), which was near to the level of statistical significance (p = 0.054).

| HRCT lobar distribution of<br>bronchiectasis | CF<br>(n=12) ‡ |  | Non – CF<br>(n=118) ‡ |  | p-value* |
| --- | --- | --- | --- | --- | --- |
|  | n | % | N | % |  |
| Right lung |  |  |  |  |  |
| Upper | 2 | 16.7% | 12 | 11.3% | 0.540 |
| Middle | 3 | 25.0% | 21 | 19.8% |  |
| Lower | 1 | 8.3% | 18 | 17.0% |  |
| Combined upper and middle | 2 | 16.7% | 22 | 20.8% |  |
| Combined upper and Lower | 0 | 0.0% | 1 | 0.9% |  |
| Combined middle and lower | 4 | 33.3% | 16 | 15.1% |  |
| Combined upper, middle and<br>lower | 0 | 0.0% | 16 | 15.1% |  |
| Total | 12 | 100.0% | 106 | 100.0% |  |
| Left lung |  |  |  |  |  |
| Upper | 3 | 27.3% | 47 | 52.8% | 0.054 |
| Lower | 2 | 18.2% | 23 | 25.8% |  |
| Combined upper and lower | 6 | 54.5% | 19 | 21.3% |  |
| Total | 11 | 100.0% | 89 | 100.0% |  |

‡ n = number
\*p-value obtained by Chi-square test, $p < 0.05$ was considered as a level of significant

## Discussion

The present study emphasizes the specific chest HRCT imaging characteristics of adult cystic fibrosis (CF) and non-CF bronchiectasis on a South Asian, tertiary care, setting. Cystic type bronchiectasis was the dominant presentation among CF patients and is the manifestation of the progressive destruction of the airways with chronic retention of mucus and repeated infection. On the other hand, non-CF patients exhibited more prevalence of cylindric type bronchiectasis, which implies relatively less severe structural distortion. These results are consistent with a study by Polverino et al^7^ and they reported that cystic changes are strongly indicative of CF even when clinical suspicion is low, but are contrary to previous study by Mason and Nakielna^14^, which indicated that cylindrical morphology is most common in CF bronchiectasis. Another study also aligns with our findings, where they reported that, in non-CF bronchiectasis had generally cylindrical, whereas in CF bronchiectasis had combination of the cylindrical and cystic morphologies^15^.

One of the most striking findings in this study was that CF was strongly correlated with a mixed pattern of central and peripheral airway involvement, which implies that CF also impacts the proximal and distal airways. Only peripheral bronchial dilatation disease was observed only in non-CF patients, so it can be used as a distinguishing feature for radiologists. This observation is consistent with pediatric research, which showed that the number of affected central and peripheral airways on inspiratory CT image was significantly greater in CF patients than control group^16^. In addition, longitudinal studies indicate that a score of peripheral bronchiectasis in CF decreases at a faster rate compared to lung function parameters, which indicates that characteristics of chest HRCT may be complementary to pulmonary function testing in measuring disease progression^13^.

Regarding lobar distribution, there was no significant difference in right lung involvement between the groups but combined upper and lower lobe involvement in left lung was more prevalent in CF. Though the correlation did not become statistically significant, the trend could be more evident in future larger cohorts. Traditionally, CF has bilateral bronchiectasis that is most frequently located in the upper lobes, whereas non-CF bronchiectasis has a more heterogenous and localized depends on underlying etiology^15^. In areas with endemic tuberculosis like Bangladesh, there is a tendency to describe upper lobe bronchiectasis as a result of a previous infection, but the presence of cystic changes with mixed central-peripheral distribution should trigger the thinking of CF and should be confirmed with an appropriate laboratory test.

There are a number of limitations in this study. The CF subgroup was comparatively small and the study was limited to one tertiary center. The retrospective nature of the study also did not allow longitudinal follow-up, which restricted the determination of the progression of the disease. Another limitation of this study was that the interpretation of the HRCT scans was done by a single radiologist. Lack of independent two-reader procedures prevented the evaluation of interobserver agreement and could have caused observer bias. Prospective, multicenter, and larger-scale studies are also required in the future in order to confirm these HRCT differences and elucidate their prognostic impact in South Asians population.

## Conclusions

Adult CF has unique HRCT patterns, such as cystic bronchiectasis, mixed airway involvement, and predominance of upper lobes, which might be used to distinguish it from non-CF bronchiectasis. It is especially important to identify these patterns in the settings where infectious etiologies are more prevalent because it can inform the earlier diagnosis and treatment of CF in adults.

## Data Availability

The data that support the findings of this study are available on request from the corresponding author. The data are not publicly available due to privacy or ethical restrictions.

## Acknowledgement

The authors gratefully acknowledge the contributions of the patients who consented to participate in this study. We extend our thanks to the clinical and laboratory staff of the Department of Respiratory Medicine, Bangladesh Medical University, for their assistance in patient management, data collection, and technical procedures.

## Authors contribution

SM was responsible for the conception and overall design of the study, development of methodology, data collection, validation, and formal statistical analysis. SM also led the investigation, and prepared the initial manuscript draft, followed by critical revision and editing. PB contributed to provide research resources, investigation, oversight, and validation of the findings, and participated in critical manuscript revision and editing. RC developed the visual representations of the data, contributed to study supervision and validation, and took part in revising and editing the manuscript. SA developed the visual representations of the data, contributed to study supervision and validation, and took part in revising and editing the manuscript. MAR coordinated project activities, supervised the research process, provided research resources, oversight, and validation of the findings, and participated in critical manuscript revision and editing. All authors reviewed the final manuscript and approved it for publication.

## Notes

### Competing Interest Statement

The authors have declared no competing interest.

### Funding Statement

Yes

### Author Declarations

This human study was approved by Institutional Review Board (IRB), Bangabandhu Sheikh Mujib Medical University, Dhaka, Bangladesh - approval: 4456

